# Factors associated with decreased anal sphincter tone and the accuracy of forced anal examinations to detect individuals having receptive anal intercourse: An observational study

**DOI:** 10.1101/2024.01.31.24302098

**Authors:** Alan G Nyitray, B R Simon Rosser, Aniruddha Hazra, Jenna Nitkowski, Derek Smith, Bridgett Brzezinski, Timothy J Ridolfi, John A Schneider, Elizabeth Y Chiao, Sandrine Sanos, Ever Mkonyi, Lucy R Mgopa, Michael W Ross

**Author notes:** Corresponding author: Dr Alan G Nyitray, Clinical Cancer Center and Center for AIDS Intervention Research Medical College of Wisconsin, 2071 North Summit Avenue Milwaukee, WI 53202.

## Abstract

**Objective and Design:** Forced anal examinations are used to prosecute sexual and gender minorities (SGM) in at least seven countries under the presumption that decreased sphincter tone, estimated by a finger inserted into the anal canal, can detect persons practicing receptive anal intercourse. In a cross-sectional analysis of the baseline data from a longitudinal study, we aimed to determine factors associated with sphincter tone and the accuracy of sphincter tonality to detect persons engaging in receptive anal intercourse.

**Setting:** Clinicians in Chicago, Houston, and Milwaukee, USA conducted digital anal rectal examinations (DARE) on 838 participants, 94.0% of whom were cisgendered males. Clinicians used the Digital Rectal Examination Scoring System to score sphincter resting tone (RT) and squeeze tone (ST). On a separate survey, individuals reported their preferred position for anal intercourse: i.e., either always/mostly insertive anal intercourse, always/mostly receptive anal intercourse, or both receptive and insertive anal intercourse. Multivariable regression assessed factors associated with decreased sphincter tone while area under the Receiver Operating Characteristic curves (AUC) estimated the accuracy of sphincter tonality to detect receptive anal intercourse.

**Results:** 11.3% had decreased RT (95/838) and 6.3% had decreased ST (53/838). The accuracy of DARE to detect any receptive anal intercourse was little better than random guessing (AUC 0.53, 95% CI 0.51 to 0.55, and AUC 0.51, 95% CI 0.49 to 0.53, respectively. RT and ST decreased with age regardless of sexual behavior (p_trend_<0.01 for both). Compared to individuals having always/mostly insertive anal intercourse, individuals having always/mostly receptive anal intercourse was associated with decreased RT, but not ST, while those equally preferring both insertive and receptive anal intercourse were not associated with decreased RT or ST.

**Conclusions:** Decreased sphincter tone is uncommon among SGM who prefer receptive anal intercourse. Given virtually no accuracy, a finger inserted into the anus has no utility to detect individuals practicing receptive anal intercourse and thus should not be used as such.

**Trial registration:** NCT04090060

**Summary Box:** *What is already known on this topic:* To gather evidence for prosecution of sexual and gender minorities, forced anal exams are used in multiple countries. The examination includes inserting the index finger into the anal canal to detect decreased sphincter tone which is considered evidence of receptive anal intercourse. We found only two small studies (n= 58 and n=24) assessing factors associated with decreased sphincter tone and none assessing the accuracy of sphincter tone to detect sexual and gender minorities having receptive anal intercourse.

*What this study adds:* Our study suggests that a finger inserted into the anal canal is not useful to detect a history of receptive anal intercourse. As such, the sexual practices of individuals cannot be known using a forced anal examination.

## Introduction

Forced anal examinations are used in at least seven countries to support prosecuting individuals suspected of being sexual minorities, gender minorities, or both.^1^ ^2^ The exam involves viewing the anus and then inserting a finger into the anal canal to assess sphincter tone. It is not related to clinical care.^3^ Reduced sphincter tone has been used in these countries as evidence that the individual engages in receptive anal intercourse (RAI) and thus is a sexual and/or gender minority. The forced anal exam result is used to support conviction of individuals for same-sex conduct, which can result in imprisonment and, in some cases, execution.^2^

Health care providers often perform the forced anal exam under the direction of police^2^ and point to a 19^th^ century text by French forensic pathologist Ambroise Tardieu, “Étude Médico-légale sur les attentats aux moeurs”^4^ to justify the use of the procedure.^2^ Tardieu’s influential studies were part of a larger medical literature interested in categorizing sexual types, especially male homosexuality or “pederasty,” which he associated with prostitution and crime,^5^ ^6^ shaping judicial response and policing of those deemed “sexual deviants” or “inverts”.^6^ ^7^ He argued that medical expertise could “locate [anatomical] evidence of pederasty” and, thus, identify those who must be prosecuted.^6^ He identified two types and claimed that, in the case of “passive pederasts”, “loose anuses,” as judged by an index finger inserted into the anal canal, indicated the individual had a history of RAI.^6^ ^8^ Tardieu also claimed that anuses of homosexual men became funnel shaped because of RAI.^4^ To our knowledge, there are no data to support Tardieu’s claims, especially the proposition that insertion of a finger into the anal canal is an accurate test of an individual engaging in RAI.

Sphincter resting tone (RT) is controlled by the internal sphincter muscle, an involuntary smooth muscle, and sphincter squeeze tone (ST) is controlled by the external sphincter muscle, a voluntary striated muscle.^9^ ^10^ Data on factors associated with RT and ST among sexual and gender minorities (SGM) come from two studies with sample sizes ≤ 58 individuals.^11^ ^12^ Sphincter tone among a large sample of SGM has not been reported to our knowledge. As part of two anal cancer screening studies among SGM that included digital anal rectal examinations (DARE) conducted by trained health care providers, we collected data on RT, ST, and sexual behavior. A DARE is a clinical procedure that includes inserting a finger into the anus to detect abnormalities and is recommended for anal cancer screening.^13^ As part of DARE, health care providers also note sphincter tone. Thus, the mechanics of the DARE clinical procedure correspond with the mechanics of the non-clinical forced anal examination. Our objective was to assess clinical and behavioral factors associated with decreased RT and ST among SGM and to estimate the accuracy of a DARE measurement of sphincter tone to detect persons who have RAI.

## Methods

### Data sources

Data for the current analyses come from two longitudinal studies assessing anal cancer screening protocols, the Prevent Anal Cancer (PAC) Self-Swab Study (R01CA215403, AGN) conducted in Milwaukee, USA^14^ and the PAC Palpation Study (R01CA232892, AGN) conducted in Chicago and Houston, USA.^15^ The studies were conducted 2020-2023. Given a common goal of anal cancer prevention and a common study population, protocols for the PAC studies were synchronized regarding recruitment, consenting procedures, clinical protocols and forms, survey procedures, and, as appropriate, survey items. All PAC studies’ procedures were approved by the Medical College of Wisconsin Human Protections Committee (PRO32999 and PRO33000). All participants in this article provided written informed consent.

### Patient and Public Involvement

Before study activities began, the PAC parent studies convened Community Advisory Boards to support participant recruitment, study activities, and data interpretation. Results from the current study have been presented to these boards.

### Participants

Individuals were eligible to join either study if they were aged ≥ 25 years and acknowledged sex with men in the past five years or identified with a minority sexual orientation or as a transgender person who has sex with men. Persons in Milwaukee were excluded if they used anticoagulants other than NSAIDS or had a prior diagnosis of anal cancer. Individuals in Chicago and Houston were excluded if they had a DARE in the prior three months, or were being treated for condyloma, hemorrhoids or anal cancer.

### Study activity and data collection

Interested persons took an online eligibility survey before being consented online. Once written consent was provided, participants took the baseline survey which had questions about sexual behavior including their preferred position during anal intercourse: “In your lifetime, have you mostly engaged in insertive anal sex, receptive anal sex, or both?” Response options were “Always or mostly engaged in insertive anal sex (your penis in his [or their] anus),” “Always or mostly engaged in receptive anal sex (his [or their] penis in your anus)” “More or less equally engaged in insertive and receptive anal sex (I have been versatile),” or “I’ve never engaged in anal sex.” Additional items asked about sexual behavior including frequency of anal intercourse and when the last anal intercourse occurred. Other items asked participants about their clinical history, including diagnoses of obesity or diabetes.

After completing the baseline survey, all individuals were asked to attend a clinic visit that included a DARE conducted by a clinician including physicians, advanced practice nurses (APN), and registered nurses. In Chicago, the clinic appointment primarily occurred at a drop-in center serving SGM. In Houston and Milwaukee, clinic appointments primarily occurred at SGM or HIV clinics. Each clinician was trained to conduct the DARE according to International Anal Neoplasia Society Guidelines and to record all perianal and anal canal abnormalities.^13^ DARE includes viewing and palpating the perianus and anal os, and then palpating the anal canal for abnormalities. As part of palpating the anal canal, sphincter tone was noted using the Digital Rectal Examination Scoring System’s (DRESS) 6-point scale for RT, 0 = “no discernible tone at rest,” 1 = “very low tone,” 2 = “mildly decreased tone,” 3 = “normal,” 4 = “elevated tone, snug” and 5 = “very high tone, a tight anal canal, difficult to insert finger” and ST, 0 = “no discernible increase in tone with squeezing effort,” 1 = “slight increase,” 2 = “fair increase but below normal,” 3 = “normal,” 4 = “strong squeeze” and 5 = “very strong squeeze, to the point of being painful to the examiner.”^16^ Clinicians did not know participants’ preferred position for anal intercourse before recording sphincter tone.

### Statistical analysis

RT was dichotomized: scores of 0-2 were defined as decreased resting tone, while scores of 3-5 were labeled as normal or increased resting tone. ST was similarly dichotomized with 0-2 defined as decreased squeeze tone and scores of 3-5 defined as normal or increased squeeze tone. For the variable preferred position for anal intercourse, the value of “never engaged in anal sex” was combined with “always or mostly engaged in insertive anal sex” (IAI).

For assessment of accuracy, individuals who mostly/always preferred RAI were combined with those who practiced RAI and IAI about equally, i.e., versatile, to construct a variable with two categories: 1) IAI and 2) those who practiced any RAI. The accuracy of decreased sphincter tone to detect preferred position for anal intercourse was assessed with 95% confidence intervals (CI) and stratified by clinician type.

Bivariate analyses for RT and ST were stratified by tonality (either decreased tone or normal/increased tone), and included assessment of their association with age, sex at birth, gender identity, race, ethnicity, sexual orientation, years of school, sexual behavior, HIV status, self-reported comorbidities (i.e., arthritis, carpal tunnel syndrome, cerebral palsy, diabetes, fibromyalgia, chronic lower back pain, motor neuron diseases, movement disorders, multiple sclerosis, obesity, spina bifida, spinal cord injury, or stroke), current anxiety and depression, clinician type, and clinical findings from DARE, BMI, and waist size.

We constructed a multivariable model to regress RT and ST on preferred position for anal intercourse and other factors after adjustment by confounders. We used purposeful modeling strategies in addition to Directed Acyclic Graphs to support identification of confounders for inclusion and intermediate variables for exclusion.^17^ ^18^ Firth bias correction was used in all regression models to reduce bias associated with cells with no data.^19^ Age, race and ethnicity, HIV status and assigned sex at birth were considered potential confounders while last RAI was an intermediate variable between preferred position for anal intercourse and sphincter tone and thus was excluded from multivariable modeling. Exposures with a likelihood ratio test p-value of less than 0.15 in bivariate analysis were included in logistic regression models and then were manually removed one at a time using backwards elimination if their p-value was > 0.05. Associations in the multivariable model were considered significant when a p-value was ≤ 0.05. Adjusted and unadjusted odds ratios were reported with 95% CIs. Study size was determined by power analyses for the primary questions in the longitudinal anal cancer studies. The current analysis did not have a separate power assessment.

Since accuracy of DARE may be related to medical training and the number of DAREs performed,^13^ supplementary analyses were conducted after restricting the dataset to physicians and APNs who performed > 50 DAREs in the study. Due to a limited number of outcomes, multivariable analysis for factors associated with sphincter ST could not be completed in this restricted dataset. Initial analyses were conducted using SAS 9.4 TS Level 1M6 (SAS Institute, Cary, North Carolina, USA) and duplicate analyses were conducted using Stata/BE 18.0 (StataCorp LLC, College Station, TX, USA) and SPSS 28.0 (Armonk, NY, USA).

## Results

A total of 1207 individuals in Chicago, Houston, and Milwaukee were eligible and gave consent to participate. Of these, 1088 completed the baseline survey including questions about their clinical history and preferred position for anal intercourse. A total of 841 individuals then attended one of nine clinics to receive a DARE and assessment of sphincter tone from a trained clinician. Three DAREs were judged as inadequate by the clinician leaving 838 individuals in the primary analysis. For supplementary analysis, i.e., restricting inclusion to individuals whose clinicians conducted > 50 DAREs in the study, 609 individuals were included in analyses.

Median age was 41 years (range, 25-80 years) and approximately one-half (52.5%) identified as non-Hispanic (NH) white, 20.9% as NH Black, and 21.1% as Hispanic (Table 1). A total of 85.3% identified as gay, and 43.5% had > 16 years of school. A total of 33.9% and 14.6% of participants self-reported a prior diagnosis of HIV and obesity, respectively. Individuals who reported IAI comprised 23.6% of the sample (n=198), 41.7% (n=349) reported preferring both receptive and insertive anal intercourse about equally, and 34.7% (n=291) reported RAI. Medical doctors, APNs, and registered nurses performed DARE on 39.5%, 52.7%, and 7.8% of participants, respectively. Clinicians noted no abnormally-shaped anuses.

**Table 1.** Characteristics of participants stratified by sphincter resting and squeeze tone, Chicago, Houston and Milwaukee, USA 2020-2022, The Prevent Anal Cancer Studies.

| Characteristic | Total<br>n=838 | Resting tone |  | p | Squeeze tone |  | p |
| --- | --- | --- | --- | --- | --- | --- | --- |
|  |  | Decreased<br>n=95 | Normal/Increased<br>n=743 |  | Decreased<br>n=53 | Normal/Increased<br>n=785 |  |
| **Sphincter tone**, mean (SD) |  | 2.97 (0.49) |  |  | 3.04 (0.48) |  |  |
| **Age**, years * median (IQR) | 41 (32-54) | 56 (49-62) | 39 (32-52) | <0.001 | 55 (47-60) | 40 (32-53) | <0.001 |
| **Body mass index** * median (IQR) | 28 (25-32) | 29 (25-35) | 28 (25-31) | 0.04 | 29 (24-33) | 28 (25-32) | 0.33 |
| **Waist size**, centimetres |  |  |  | <0.001 |  |  | 0.003 |
| ≤ 102 | 534 (64.3) | 42 (44.2) | 492 (66.9) |  | 24 (45.3) | 510 (65.6) |  |
| > 102 | 297 (35.7) | 53 (55.8) | 244 (33.2) |  | 29 (54.7) | 268 (34.5) |  |
| **Assigned sex at birth** † |  |  |  | 0.007 |  |  | 0.04 |
| Male | 826 (98.6) | 90 (94.7) | 736 (99.1) |  | 50 (94.3) | 776 (98.9) |  |
| Female | 12 (1.4) | 5 (5.3) | 7 (0.9) |  | 3 (5.7) | 9 (1.2) |  |
| **Gender identity** † |  |  |  | 0.002 |  |  | 0.17 |
| Man | 787 (94.0) | 86 (90.5) | 701 (94.5) |  | 48 (90.6) | 739 (94.3) |  |
| Transgender woman, transgender man, woman, and other | 29 (3.5) | 9 (9.5) | 20 (2.7) |  | 4 (7.6) | 25 (3.2) |  |
| Non-binary | 21 (2.5) | 0 | 21 (2.8) |  | 1 (1.9) | 20 (2.6) |  |
| **Race and ethnicity** |  |  |  | 0.20 |  |  | 0.05 |
| White, non-Hispanic | 437 (52.5) | 56 (59.0) | 381 (51.6) |  | 36 (67.9) | 401 (51.4) |  |
| Black, non-Hispanic | 174 (20.9) | 18 (19.0) | 156 (21.1) |  | 10 (18.9) | 164 (21.0) |  |
| Hispanic | 176 (21.1) | 18 (19.0) | 158 (21.4) |  | 5 (9.4) | 171 (21.9) |  |
| Asian, non-Hispanic | 30 (3.6) | 0 | 30 (4.1) |  | 0 | 30 (3.9) |  |
| Other, non-Hispanic ‡ | 16 (1.9) | 3 (3.2) | 13 (1.8) |  | 2 (3.8) | 14 (1.8) |  |
| **Sexual orientation** † |  |  |  | 0.24 |  |  | 0.64 |
| Gay | 713 (85.3) | 86 (90.5) | 627 (84.6) |  | 46 (86.8) | 667 (85.2) |  |
| Bisexual | 73 (8.7) | 4 (4.2) | 69 (9.3) |  | 3 (5.7) | 70 (8.9) |  |
| Queer | 39 (4.7) | 3 (3.2) | 36 (4.9) |  | 3 (5.7) | 36 (4.6) |  |
| Heterosexual, lesbian, other, or don't know ** | 11 (1.3) | 2 (2.1) | 9 (1.2) |  | 1 (1.9) | 10 (1.3) |  |
| **Years of school** □ □ |  |  |  | 0.03 |  |  | 0.23 |
| ≤ 12 | 96 (11.5) | 14 (14.7) | 82 (11.1) |  | 8 (15.1) | 88 (11.3) |  |
| 13-15 | 186 (22.4) | 30 (31.6) | 156 (21.2) |  | 14 (26.4) | 172 (22.1) |  |
| 16 | 188 (22.6) | 17 (17.9) | 171 (23.2) |  | 11 (20.8) | 177 (22.7) |  |
| >16 | 362 (43.5) | 34 (35.8) | 328 (44.5) |  | 20 (37.7) | 342 (43.9) |  |
| **Marital status** |  |  |  | 0.05 |  |  | 0.14 |
| Single no steady partner | 365 (44.1) | 38 (40.9) | 327 (44.6) |  | 23 (43.4) | 342 (44.2) |  |
| Married | 155 (18.7) | 14 (15.1) | 141 (19.2) |  | 5 (9.4) | 150 (19.4) |  |
| Single with steady partner(s) | 146 (17.7) | 15 (16.1) | 131 (17.9) |  | 12 (22.6) | 134 (17.3) |  |
| Cohabiting | 105 (12.7) | 13 (14.0) | 92 (12.5) |  | 6 (11.3) | 99 (12.8) |  |
| Widowed, divorced, or separated | 56 (6.8) | 13 (14.0) | 43 (5.9) |  | 7 (13.2) | 49 (6.3) |  |
| **Any anal canal abnormality detected at visit** |  |  |  | 0.31 |  |  | 0.02 |
| No | 725 (86.5) | 79 (83.2) | 646 (86.9) |  | 40 (75.5) | 685 (87.3) |  |
| Yes | 113 (13.5) | 16 (16.8) | 97 (13.1) |  | 13 (24.5) | 100 (12.7) |  |
| **Anal condyloma detected at visit <sup>†</sup>** |  |  |  | 0.17 |  |  | 0.04 |
| No | 825 (98.5) | 92 (96.8) | 733 (98.7) |  | 50 (94.3) | 775 (98.7) |  |
| Yes | 13 (1.5) | 3 (3.2) | 10 (1.3) |  | 3 (5.7) | 10 (1.3) |  |
| **HIV diagnosis history** |  |  |  | <0.001 |  |  | 0.99 |
| No | 548 (66.1) | 42 (44.2) | 506 (68.9) |  | 35 (66.0) | 513 (66.1) |  |
| Yes | 281 (33.9) | 53 (55.8) | 228 (31.1) |  | 18 (34.0) | 263 (33.9) |  |
| **Diabetes diagnosis history** |  |  |  | <0.001 |  |  | 0.01<sup>†</sup> |
| No | 775 (92.5) | 77 (81.1) | 698 (93.9) |  | 44 (83.0) | 731 (93.1) |  |
| Yes | 63 (7.5) | 18 (18.9) | 45 (6.1) |  | 9 (17.0) | 54 (6.9) |  |
| **Obesity diagnosis history** |  |  |  | 0.002 |  |  | 0.61 |
| No | 716 (85.4) | 71 (74.7) | 645 (86.8) |  | 44 (83.0) | 672 (85.6) |  |
| Yes | 122 (14.6) | 24 (25.3) | 98 (13.2) |  | 9 (17.0) | 113 (14.4) |  |
| **Arthritis diagnosis history** |  |  |  | <0.001 |  |  | 0.05 |
| No | 748 (89.3) | 74 (77.9) | 674 (90.7) |  | 43 (81.1) | 705 (89.8) |  |
| Yes | 90 (10.7) | 21 (22.1) | 69 (9.3) |  | 10 (18.9) | 80 (10.2) |  |
| **Stroke diagnosis history <sup>†</sup>** |  |  |  | 0.04 |  |  | 0.55 |
| No | 826 (98.6) | 91 (95.8) | 735 (98.9) |  | 52 (98.1) | 774 (98.6) |  |
| Yes | 12 (1.4) | 4 (4.2) | 8 (1.1) |  | 1 (1.9) | 11 (1.4) |  |
| **Preferred position for anal intercourse** |  |  |  | 0.06 |  |  | 0.69 |
| Always/mostly insertive | 198 (23.6) | 14 (14.7) | 184 (24.8) |  | 10 (18.9) | 188 (24.0) |  |
| Versatile | 349 (41.7) | 40 (42.1) | 309 (41.6) |  | 23 (43.4) | 326 (41.5) |  |
| Always/mostly receptive | 291 (34.7) | 41 (43.2) | 250 (33.7) |  | 20 (37.7) | 271 (34.5) |  |
| **Frequency of anal intercourse in the last 30 days <sup>□□</sup>** |  |  |  | 0.30 |  |  | 0.02 |
| 0 | 294 (35.6) | 41 (43.6) | 253 (34.6) |  | 30 (56.6) | 264 (34.2) |  |
| 1 | 117 (14.2) | 14 (14.9) | 103 (14.1) |  | 6 (11.3) | 111 (14.4) |  |
| 2-3 | 188 (22.8) | 12 (12.8) | 176 (24.0) |  | 6 (11.3) | 182 (23.5) |  |
| 1 per week | 122 (14.8) | 14 (14.9) | 108 (14.8) |  | 3 (5.7) | 119 (15.4) |  |
| > 1 per week | 105 (12.7) | 13 (13.8) | 92 (12.6) |  | 8 (15.1) | 97 (12.6) |  |
| **Last receptive anal intercourse <sup>†</sup>** |  |  |  | 0.04 |  |  | 0.02 |
| Within last month | 364 (43.8) | 39 (41.1) | 325 (44.1) |  | 15 (28.3) | 349 (44.8) |  |
| > 1 month | 394 (47.4) | 45 (47.4) | 349 (47.4) |  | 34 (64.2) | 360 (46.2) |  |
| Never | 33 (4.0) | 1 (1.1) | 32 (4.3) |  | 0 | 33 (4.2) |  |
| Don't know | 41 (4.9) | 10 (10.5) | 31 (4.2) |  | 4 (7.6) | 37 (4.8) |  |
| **Clinician type** |  |  |  | <0.001 |  |  | <0.001 |
| Medical doctor | 331 (39.5) | 12 (12.6) | 319 (42.9) |  | 8 (15.1) | 323 (41.2) |  |
| Advanced practice nurse | 442 (52.7) | 69 (72.6) | 373 (50.2) |  | 25 (47.2) | 417 (53.1) |  |
| Registered nurse | 65 (7.8) | 14 (14.7) | 51 (6.9) |  | 20 (37.7) | 45 (5.7) |  |
| Data are n (%). Missing: Waist size n = 7; gender identity n = 1; race and ethnicity n = 5; sexual orientation n = 2; years of school n = 6; marital n = 11; HIV diagnosis history n = 9; frequency of anal intercourse in the last 30 days n = 12; last receptive anal intercourse n = 6. Abbreviations: SD, standard deviation; IQR, interquartile range. All hypothesis tests are Pearson chi square unless otherwise noted. |  |  |  |  |  |  |  |
| * Student's *t*-test |  |  |  |  |  |  |  |
| † Fisher's Exact Test |  |  |  |  |  |  |  |
| ‡ Other includes Native Hawaiian or Pacific Islander, American Indian or Alaskan Native, and Other. |  |  |  |  |  |  |  |
| ** Individuals' self-reported sexual orientation included "heterosexual" (n=1) and "lesbian" (n=2) although self-reported sexual behavior satisfied inclusion criteria. |  |  |  |  |  |  |  |
| □□ Cochran-Armitage Test for Trend |  |  |  |  |  |  |  |

### Sphincter resting tone

Clinicians identified 95 (11.3%) individuals with decreased RT and 743 (90.3%) with normal or increased RT (Table 1). On the 0-5 DRESS scale, the mean RT was 2.97 (SD, 0.49) (Table 1). The mean RT among IAI, versatile, and RAI individuals was 3.10 (SD, 0.54), 2.95 (SD, 0.47), and 2.91 (0.47), respectively (p < 0.001; data not shown).

The median age of persons with decreased RT was 56 years while the age of persons with normal or increased RT was 39 years (p<0.001) (Table 1). Regardless of preferred position for anal intercourse, increased age was associated with decreased RT (Figure).

**Figure.**
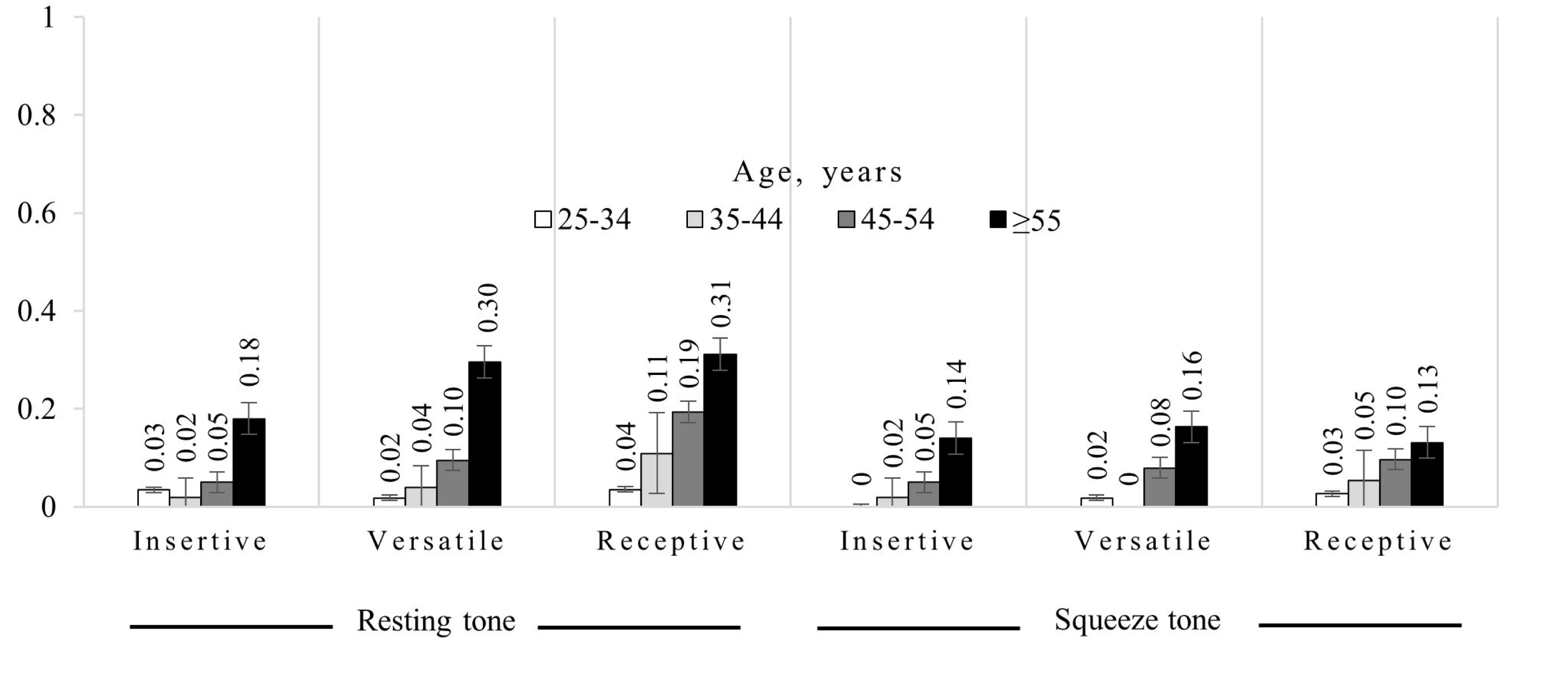
The proportion of individuals with decreased sphincter resting and squeeze tone by age and preferred position for anal intercourse in the Prevent Anal Cancer Studies, Chicago, Houston, and Milwaukee, USA, 2020-2022. The Cochrane-Armitage test for trend p-value is <0.01 for each histogram.

Individuals with decreased RT also reported obesity almost twice as often as those with normal or increased tone (p=0.002) (Table 1). A higher proportion of people with HIV had decreased RT compared with HIV-negative persons (18.9% and 7.7%, respectively, p < 0.001) and individuals reporting a diagnosis of diabetes or arthritis also had decreased RT (p < 0.001, Table 1).

In univariate analysis (Table 2), increasing age, BMI, and waist size were associated with decreased RT. Assigned sex at birth, gender identity, marital status, HIV status, diabetes, obesity, arthritis, stroke, preferred position for anal intercourse, frequency of any anal intercourse, and clinician type were also associated with RT. For example, reporting a prior diagnosis of obesity was associated with decreased RT (OR 2.25, 95% CI 1.35-3.73, compared to those not reporting obesity). Individuals preferring RAI were associated with decreased RT (OR 2.11, 95% CI 1.12-2.95 compared to individuals preferring IAI).

**Table 2:** Factors associated with sphincter resting and squeeze tone, Chicago, Houston, and Milwaukee, USA 2020-2022.

|  | Resting tone |  | Squeeze tone |  |
| --- | --- | --- | --- | --- |
|  | OR (95% CI) | aOR* (95% CI) | OR (95% CI) | aOR† (95% CI) |
| **Age, years** | **1.08 (1.06-1.10)** | **1.09 (1.06-1.11)** | **1.07 (1.05-1.10)** | **1.07 (1.04-1.09)** |
| **Body mass index** | **1.03 (1.00-1.06)** | - | - | - |
| **Waist size, centimetres** |  |  |  |  |
| ≤ 102 | 1.0 |  | 1.0 | - |
| > 102 | **2.54 (1.65-3.90)** | - | **2.29 (1.31-3.99)** | - |
| **Assigned sex at birth** |  |  |  |  |
| Male | 1.0 | - | 1.0 | 1.0 |
| Female | **5.97 (1.86-19.15)** | - | **5.66 (1.53-20.93)** | **13.84 (3.19-60.05)** |
| **Gender identity** |  |  |  |  |
| Man | 1.0 | 1.0 | - | - |
| Transgender woman, transgender man, woman, and other | **3.76 (1.67-8.48)** | **3.55 (1.11-11.38)** | - | - |
| Non-binary | 0.19 (0.01-3.36) | 0.45 (0.02-9.04) | - | - |
| **Race and ethnicity** |  |  |  |  |
| White, non-Hispanic | 1.0 | 1.0 | 1.0 | - |
| Black, non-Hispanic | 0.80 (0.46-1.40) | 1.62 (0.82-3.22) | 0.70 (0.35-1.43) | - |
| Hispanic | 0.79 (0.45-1.38) | 1.37 (0.71-2.63) | **0.35 (0.14-0.88)** | - |
| Asian, non-Hispanic | 0.11 (0.01-1.92) | 0.16 (0.01-3.67) | 0.18 (0.01-3.15) | - |
| Other, non-Hispanic □ | 1.75 (0.51-6.07) | **6.06 (1.37-26.83)** | 1.90 (0.46-7.88) | - |
| **Years of school** |  |  |  |  |
| ≤ 12 | 1.0 |  | - | - |
| 13-15 | 1.11 (0.56-2.20) | - | - | - |
| 16 | 0.58 (0.28-1.23) | - | - | - |
| >16 | 0.60 (0.31-1.16) | - | - | - |
| **Marital status** |  |  |  |  |
| Single no steady partner | 1.0 |  | 1.0 | 1.0 |
| Married | 0.87 (0.46-1.65) | - | 0.53 (0.21-1.38) | **0.35 (0.13-0.94)** |
| Single with steady partner(s) | 1.00 (0.54-1.87) | - | 1.36 (0.66-2.77) | 1.28 (0.57-2.87) |
| Cohabiting | 1.24 (0.64-2.41) | - | 0.95 (0.39-2.34) | 0.75 (0.28-2.01) |
| Widowed, divorced, or separated | **2.64 (1.31-5.32)** | - | 2.21 (0.92-5.33) | 1.01 (0.38-2.66) |
| **Anal condyloma detected at visit** |  |  |  |  |
| No | - | - | 1.0 | 1.0 |
| Yes | - | - | **5.12 (1.41-18.55)** | **6.59 (1.42-30.59)** |
| **HIV diagnosis history** |  |  |  |  |
| No | 1.0 | 1.0 | - | - |
| Yes | **2.79 (1.81-4.30)** | **1.72 (1.01-2.92)** | - | - |
| **Diabetes diagnosis history** |  |  |  |  |
| No | 1.0 | - | 1.0 | - |
| Yes | **3.66 (2.02-6.63)** | - | **2.87 (1.34-6.11)** | - |
| **Obesity diagnosis history** |  |  |  |  |
| No | 1.0 | 1.0 |  | - |
| Yes | **2.25 (1.35-3.73)** | **2.10 (1.17-3.79)** |  | - |
| **Arthritis diagnosis history** |  |  |  |  |
| No | 1.0 | - | 1.0 | - |
| Yes | **2.80 (1.63-4.82)** | - | **2.12 (1.03-4.33)** | - |
| **Stroke diagnosis history** |  |  |  |  |
| No | 1.0 | - | - | - |
| Yes | **4.26 (1.27-14.26)** | - | - | - |
| **Preferred position for anal** |  |  |  |  |
| **intercourse** |  |  |  |  |
| Always/mostly insertive | 1.0 | 1.0 | 1.0 | - |
| Versatile | 1.67 (0.89-3.12) | 1.80 (0.89-3.66) | 1.29 (0.61-2.74) | - |
| Always/mostly receptive | **2.11 (1.12-2.95)** | **2.82 (1.37-5.82)** | 1.36 (0.63-2.92) | - |
| **Frequency of anal intercourse in the last 30 days** |  |  |  |  |
| 0 | 1.0 |  | 1.0 | - |
| 1 | 0.86 (0.45-1.63) | - | 0.51 (0.21-1.22) | - |
| 2-3 | **0.43 (0.22-0.84)** | - | 0.31 (0.13-0.74) | - |
| 1 per week | 0.82 (0.43-1.55) | - | **0.25 (0.08-0.79)** | - |
| > 1 per week | 0.89 (0.46-1.73) | - | 0.76 (0.34-1.68) | - |
| **Clinician type** |  |  |  |  |
| Medical doctor | 1.0 | 1.0 | 1.0 | 1.0 |
| Advanced practice nurse | **4.76 (2.56-8.85)** | **3.51 (1.77-6.94)** | **2.32 (1.05-5.13)** | 1.93 (0.87-4.32) |
| Registered nurse | **7.20 (3.18-16.27)** | **5.91 (2.38-14.69)** | **17.15 (7.25-40.55)** | **11.79 (4.71-29.52)** |
| Before rounding to two decimals, bolded point estimates and 95% CI did not include unity. Abbreviations: OR, odds ratio; aOR, adjusted odds ratio; CI, confidence interval. |  |  |  |  |
| * Adjusted by variables retained in model and potential confounder assigned sex at birth. |  |  |  |  |
| † Adjusted by variables retained in model and potential confounder race and ethnicity. |  |  |  |  |
| □ Other includes Native Hawaiian or Pacific Islander, American Indian or Alaskan Native, and other. |  |  |  |  |

In multivariable analysis for decreasing RT, age, gender identity, race, HIV status, obesity, and clinician type were significant while preferred position for anal intercourse was also associated with RT after adjusting for confounders (aOR 2.82, 95% CI 1.37-5.82 for persons who performed RAI compared to those performing IAI) (Table 2). Engaging equally in RAI and IAI was not associated with decreased RT.

In supplementary analysis with clinicians completing DAREs with > 50 individuals in the study, a total of 609 individuals were analyzed (Supplementary Table 1) with the prevalence of decreased RT being 7.4%. In multivariable analysis in this restricted dataset, factors associated with decreased RT were similar to the full dataset: increasing age, obesity, preferring always/mostly RAI (compared to IAI), and an APN clinician type (compared to physician); however, gender identity and race/ethnicity were no longer associated with decreased RT.

### Sphincter squeeze tone

Clinicians detected 6.3% (n=53) of individuals with decreased ST (Table 1). On the 0-5 DRESS scale, the mean ST was 3.04 (SD, 0.48). By preferred position for anal intercourse, the mean DRESS scale score was 3.10 (SD, 0.55), 3.02 (SD, 0.45), and 3.01 (SD, 0.44) among individuals preferring IAI, being versatile, and RAI, respectively (p = 0.12) (data not shown).

As with RT, the median age of persons with decreased ST was higher than those with normal/increased tone, 55 and 40 years, respectively (p<0.001, Table 1). Increasing age was associated with decreased ST regardless of preferred position for anal intercourse (p<0.01, figure). Unlike with RT, there was no difference in ST by HIV status (p<0.001 and p=0.99, respectively). Also unlike RT, there was little difference in ST among people with and without a self-reported diagnosis of obesity (p=0.002 and p=0.61).

In univariate logistic regression analysis for ST (Table 2), significant associations between factors and ST were generally similar to associations between factors and RT, for example, age, waist size, assigned sex at birth, diabetes, arthritis, frequency of anal intercourse and clinician type. Unlike with RT, BMI, gender identity, marital status, HIV status, obesity, stroke, and preferred position for anal sex were not associated with ST while race/ethnicity and condyloma detected at the clinic visit were associated with ST.

In multivariable analysis (Table 2), decreased ST was associated with increasing age, being assigned female sex at birth (compared to being assigned male), marital status, a condyloma diagnosis at the clinic visit, and clinician type of registered nurse (compared with physician) after adjusting for other variables, including confounders, in the model. Being married, compared to single with no steady partner(s), was protective of decreased ST. Individuals preferring RAI or being versatile were not significantly associated with decreased ST.

### Accuracy

The AUC of a DARE assessment of RT to detect individuals having any RAI was 0.53 (95% CI 0.51-0.55) (Table 3). The accuracy of a DARE assessment of ST to detect individuals having any RAI was 0.51 (95% CI 0.49-0.53).

**Table 3:**
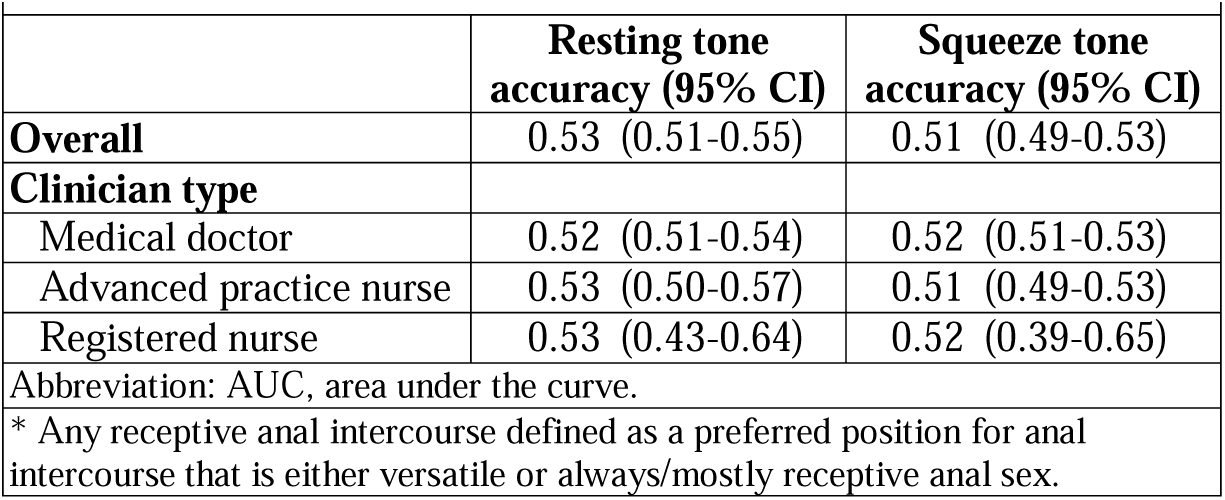
Accuracy of digital anal rectal examinations to detect persons engaging in any receptive anal intercourse,* Chicago, Houston, and Milwaukee, USA 2020-2022.

In supplementary analyses with only physicians and APNs, the accuracy of a DARE assessment of RT to detect individuals having any RAI was 0.09 (95% CI 0.07-0.12) (Supplementary Table 3). The accuracy of a DARE assessment of ST to detect individuals having any RAI was 0.03 (95% CI 0.02-0.05).

## Discussion

We report data that rejects the utility of the forced anal exam to detect persons having receptive anal intercourse. In this sample of 838 individuals who were sexual minorities, gender minorities, or both, decreased anal sphincter resting tone was uncommon (11.3%) with 88.7% of individuals having normal or increased RT based on a DARE DRESS score. For squeeze tone, 93.7% of SGM had normal or increased ST. The accuracy of a DARE assessment of RT and ST to detect individuals having any RAI was only 0.53 and 0.51, respectively. Since an AUC value of 0.50 means a test is no better than flipping a coin, the accuracy of sphincter tonality to detect receptive anal intercourse is virtually nil. A limitation of this finding is that sphincter tone may differ for individuals experiencing forced vs. voluntary examinations. If forced exams result in tensing or sphincter contraction individuals, regardless of preferred position for anal sex, we would expect accuracy of the exam to decrease.

Decreased anal sphincter RT and ST was strongly associated with age in all categories of preferred position for anal intercourse. With each year of age, there was a 9% increased risk of having decreased RT. Age-associated decreased sphincter tone has ample support in the literature.^20–22^ Age-related fibrotic processes likely affect both smooth^23^ and striated^10^ muscles, reducing muscle content and enhancing atrophy;^9^ ^24^ thus, in this sample, age-related exposures, i.e., diabetes, arthritis, and stroke, were associated with decreased tonality in univariate analysis but not after adjusting for age.

Large studies have assessed sphincter tone with manometry,^20^ ^25^ ^26^ a device to measure sphincter pressure, but none assessed sphincter tone among SGM. Two small studies^11^ ^12^ have used manometry to assess the association between sphincter tone and RAI. Miles et al.^11^ compared 40 gay males having RAI (23 of whom had HIV) with 18 age-matched heterosexual males who did not acknowledge RAI. Chun et al.^12^ enrolled 14 HIV-negative gay males having RAI and ten age-matched male controls not acknowledging RAI. Both studies observed lower RT among the gay males than the controls, while ST did not differ between the two groups. These results are consistent with our observations. Chun et al. suggested that lower RT pressures during manometry may occur if the internal sphincter muscle of those preferring RAI is better able to relax during anal canal manometry compared to those acknowledging only IAI.

While these data indicate that SGM seldom have decreased sphincter tone (only 14.1% of those preferring RAI had decreased RT), they also suggest an association with decreased resting tone for those always/mostly having RAI but no significant association with those having *any* RAI, i.e., those preferring RAI and those equally preferring both RAI and IAI. The strongest influence on tone was age, regardless of preferred position for anal intercourse. Note that for those preferring RAI, age-related decreased sphincter tone may result in decreased anodyspareunia.^27^ ^28^

In general, factors associated with decreased RT and ST were similar with notable exceptions including those always/mostly preferring RAI. Perhaps individuals preferring RAI were better able to relax the external sphincter muscle during the DARE than individuals preferring IAI. Individuals with HIV were also associated with decreased RT but not ST which may be related to HIV-induced accelerated aging and damage to smooth muscle cells caused by HIV and anti-retroviral treatment.^29^

Those with decreased RT and ST had increased odds of receiving a DARE from an APN or registered nurse compared to a physician. These results were consistent with the restricted data set which used observations from clinicians conducting >50 DAREs in the study. In Houston, where APNs completed all DAREs, participants were substantially older and more likely to have HIV compared to Chicago participants where most DAREs were conducted by a physician; thus, residual age- or HIV-related confounding may explain differences in sphincter tone by clinician type. Alternatively, differences in tone assessment might occur if physicians receive more digital rectal examination training or practice in school than APNs or registered nurses.

To our knowledge, the association between obesity and sphincter resting tone has not been assessed. Obesity is associated with chronic inflammation^30^ which may induce fibrotic processes, in turn, promoting internal sphincter atrophy.^20^

The combined category of transgender, woman, and other had decreased resting sphincter tone although data were sparse. Just over one-quarter (27.6%), reported a female assignment at birth. Lower RT in females than males may be due to parity and increased muscle mass in males.^20–22^

### Limitations

Practical limitations in this large study required using the DRESS scale as a proxy for manometry. While the DRESS scale is strongly correlated with manometry pressures (Spearman rank correlation, 0.82 for RT and 0.81 for ST),^16^ it may introduce more error in sphincter tone outcome scores. Nonetheless, previous manometry findings associating decreased sphincter tone with age, sex at birth, and RAI are consistent with the current study’s findings. Two clinicians, a physician and APN, conducted 65% of DAREs which may increase reliability in resting and squeeze sphincter tone measurement. Self-reported lifetime preferred position for anal intercourse, disease history, and frequency of anal intercourse is subject to recall bias. No data were collected about douching, sex toys, or other anal manipulations which may limit our understanding of factors associated with sphincter tone. The sample was diverse regarding age, race, ethnicity, and gender identity; however, it was also highly educated with 43.5% reporting > 16 years of school.

In this study with 838 SGM, decreased sphincter resting tone and squeeze tone were rare. DARE had almost no accuracy to detect individuals having RAI; thus, the forced anal examination is useless in identifying SGM who prefer RAI, while also causing significant harm to individuals subjected to the test.^1–3^ The proposition that a finger inserted into the anal canal can detect an individual engaging in RAI is false. Increased age was consistently associated with decreased sphincter resting tone and squeeze tone.

## Data Availability

Fully de-identified datasets and data dictionary will be shared with properly trained investigators requesting such on the study website (https://mindyourbehind.org) within one year of study completion after assessment of institutional policies, Medical College of Wisconsin Human Research Protections Program rules, as well as local, state, and federal and tribal laws and regulations. Further information is available from the corresponding author upon request.

## Author Contributions

AGN, SS, EYC, BRSR, and MWR conceptualized the analysis and developed the methodology. AH and DS conducted a majority of clinical examinations including DARE. AGN managed the data. AGN and JN conducted the formal analysis. AGN wrote the first draft of the manuscript. AGN, BRSR, AH, JN, DS, AP, BB, TJR, JAS, EYC, SS, EM, LRM, and MWR reviewed and edited manuscript drafts. The corresponding author attests that all listed authors meet authorship criteria and that no others meeting the criteria have been omitted.

*“The Corresponding Author has the right to grant on behalf of all authors and does grant on behalf of all authors, a worldwide licence to the Publishers and its licensees in perpetuity, in all forms, formats and media (whether known now or created in the future), to i) publish, reproduce, distribute, display and store the Contribution, ii) translate the Contribution into other languages, create adaptations, reprints, include within collections and create summaries, extracts and/or, abstracts of the Contribution, iii) create any other derivative work(s) based on the Contribution, iv) to exploit all subsidiary rights in the Contribution, v) the inclusion of electronic links from the Contribution to third party material where-ever it may be located; and, vi) licence any third party to do any or all of the above.”*

## Acknowledgements

We thank the participants in the studies, the Prevent Anal Cancer (PAC) Self-Swab and Palpation Study Community Advisory Boards, the PAC Self-Swab and Palpation Study Teams, and Dr Jason S Farr.

## Funding and Disclaimer

This research was supported by the National Cancer Institute of the National Institutes of Health under award numbers R01CA215403 and R01CA232892. The content is solely the responsibility of the authors and does not necessarily represent the official views of the National Institutes of Health. This work was also supported by the Medical College of Wisconsin. The study sponsors had no role in study design; in the collection, analysis, and interpretation of data; in the writing of the report; or in the decision to submit the paper for publication. The study authors had full access to the data and take responsibility for its integrity and accuracy.

## Transparency declaration

The lead author (AGN) affirms that the manuscript is an honest, accurate, and transparent account of the study being reported; that no important aspects of the study have been omitted; and that any discrepancies from the study as originally planned (and, if relevant, registered) have been explained.

