## Supplemental Material for "Factors associated with decreased anal sphincter tone and the accuracy of forced anal examinations to detect individuals having receptive anal intercourse: An observational study"

**Supplementary Materials**

**Table of Contents**

**Supplemental Table 1. Characteristics of participants stratified by sphincter resting and squeeze tone measured by physicians and advanced practice nurses completing >50 digital anal rectal examinations, Chicago and Houston, USA 2020-2022, The Prevent Anal Cancer Studies**

**Supplemental Table 2*:* Factors associated with sphincter resting tone measured by physicians and advanced practice nurses completing >50 digital anal rectal examinations, Chicago and Houston, USA 2020-2022**

**Supplemental Table 3*:* Accuracy of digital anal rectal examinations to detect persons engaging in any receptive anal intercourse* as measured by physicians and advanced practice nurses completing >50 examinations, Chicago, and Houston USA 2020-2022**

| **Supplemental Table 1. Characteristics of participants stratified by sphincter resting and squeeze tone measured by physicians and advanced practice nurses completing >50 digital anal rectal examinations, Chicago and Houston, USA 2020-2022, The Prevent Anal Cancer Studies** | | | | | | | |
| --- | --- | --- | --- | --- | --- | --- | --- |
|  | | **Resting tone** | |  | **Squeeze tone** | |  |
| **Characteristic** | **Total**  **n=609** | **Decreased**  **n=51** | **Normal/Increased**  **n=558** | **p** | **Decreased**  **n=16** | **Normal/Increased**  **n=593** | **p** |
| **Sphincter tone**, mean (SD) |  | 2.98 (0.43) | |  | 3.04 (0.35) | |  |
| **Age**, years * median (IQR) | 39 (32-52) | 55 (47-64) | 38 (31-51) | <0.001 | 56 (51-65) | 39 (32-52) | <0.001 |
| **Body mass index** * median (IQR) | 28 (25-32) | 29 (25-35) | 28 (25-31) | 0.14 | 26 (24-35) | 28 (25-32) | 0.99 |
| **Waist size**, centimetres |  |  |  | 0.003 |  |  | 0.13 |
| ≤ 102 | 409 (67.6) | 25 (49.0) | 384 (69.3) |  | 8 (50.0) | 401 (68.1) |  |
| > 102 | 196 (32.4) | 26 (51.0) | 170 (30.7) |  | 8 (50.0) | 188 (31.9) |  |
| **Assigned sex at birth** ^†^ |  |  |  | 0.41 |  |  | 0.99 |
| Male | 603 (99.0) | 50 (98.0) | 553 (99.1) |  | 16 (100.0) | 587 (99.0) |  |
| Female | 6 (1.0) | 1 (2.0) | 5 (0.9) |  | 0 | 6 (1.0) |  |
| **Gender identity** ^†^ |  |  |  | 0.19 |  |  | 0.20 |
| Man | 576 (94.7) | 48 (94.1) | 528 (94.8) |  | 14 (87.5) | 562 (94.9) |  |
| Transgender woman, transgender man, woman, and other | 16 (2.6) | 3 (5.9) | 13 (2.3) |  | 1 (6.3) | 15 (2.5) |  |
| Non-binary | 16 (2.6) | 0 | 16 (2.9) |  | 1 (6.3) | 15 (2.5) |  |
| **Race and ethnicity** |  |  |  | 0.23 |  |  | 0.23^†^ |
| White, non-Hispanic | 298 (49.3) | 31 (60.8) | 267 (48.2) |  | 9 (56.3) | 289 (49.1) |  |
| Black, non-Hispanic | 128 (21.2) | 9 (17.7) | 119 (21.5) |  | 5 (31.3) | 123 (20.9) |  |
| Hispanic | 137 (22.6) | 9 (17.7) | 128 (23.1) |  | 1 (6.3) | 136 (23.1) |  |
| Asian, non-Hispanic | 28 (4.6) | 0 | 28 (5.1) |  | 0 | 28 (4.8) |  |
| Other, non-Hispanic ^‡^ | 14 (2.3) | 2 (3.9) | 12 (2.2) |  | 1 (6.3) | 13 (2.2) |  |
| **Sexual orientation** ^†^ |  |  |  | 0.20 |  |  | 0.75 |
| Gay | 526 (86.5) | 48 (94.1) | 478 (85.8) |  | 16 (100.0) | 510 (86.2) |  |
| Bisexual | 45 (7.4) | 2 (3.9) | 43 (7.7) |  | 0 | 45 (7.6) |  |
| Queer | 29 (4.8) | 0 | 29 (5.2) |  | 0 | 29 (4.9) |  |
| Lesbian, other, or don’t know ** | 8 (1.3) | 1 (2.0) | 7 (1.3) |  | 0 | 8 (1.4) |  |
| **Years of school** ^⸹⸹^ |  |  |  | 0.11 |  |  | 0.34 |
| ≤ 12 | 62 (10.3) | 7 (13.7) | 55 (10.0) |  | 3 (18.8) | 59 (10.0) |  |
| 13-15 | 134 (22.2) | 16 (31.4) | 118 (21.3) |  | 4 (25.0) | 130 (22.1) |  |
| 16 | 137 (22.7) | 9 (17.7) | 128 (23.2) |  | 2 (12.5) | 135 (23.0) |  |
| >16 | 271 (44.9) | 19 (37.3) | 252 (45.6) |  | 7 (43.8) | 264 (44.9) |  |
| **Marital status** |  |  |  | 0.37 |  |  | 0.83 ^†^ |
| Single no steady partner | 268 (44.5) | 20 (39.2) | 248 (45.0) |  | 8 (50.0) | 260 (44.4) |  |
| Married | 121 (20.1) | 11 (21.6) | 110 (20.0) |  | 2 (12.5) | 119 (20.3) |  |
| Single with steady partner(s) | 108 (17.9) | 8 (15.7) | 100 (18.2) |  | 4 (25.0) | 104 (17.8) |  |
| Cohabitating | 71 (11.8) | 6 (11.8) | 65 (11.8) |  | 1 (6.3) | 70 (12.0) |  |
| Widowed, divorced, or separated | 34 (5.7) | 6 (11.8) | 28 (5.1) |  | 1 (6.3) | 33 (5.6) |  |
| **Any anal canal abnormality detected at visit** |  |  |  | 0.50 |  |  | 0.07^†^ |
| No | 521 (85.6) | 42 (82.4) | 479 (85.8) |  | 11 (68.8) | 510 (86.0) |  |
| Yes | 88 (14.5) | 9 (17.7) | 79 (14.2) |  | 5 (31.3) | 83 (14.0) |  |
| **Anal condyloma detected at visit** ^†^ |  |  |  | 0.46 |  |  | 0.17 |
| No | 602 (98.9) | 50 (98.0) | 552 (98.9) |  | 15 (93.8) | 587 (99.0) |  |
| Yes | 7 (1.2) | 1 (2.0) | 6 (1.1) |  | 1 (6.3) | 6 (1.0) |  |
| **HIV diagnosis history** |  |  |  | 0.01 |  |  | 0.17 |
| No | 396 (65.9) | 25 (49.0) | 371 (67.5) |  | 8 (50.0) | 388 (66.3) |  |
| Yes | 205 (34.1) | 26 (51.0) | 179 (32.6) |  | 8 (50.0) | 197 (33.7) |  |
| **Diabetes diagnosis history** ^†^ |  |  |  | 0.01 |  |  | 0.08 |
| No | 570 (93.6) | 43 (84.3) | 527 (94.4) |  | 13 (81.3) | 557 (93.9) |  |
| Yes | 39 (6.4) | 8 (15.7) | 31 (5.6) |  | 3 (18.8) | 36 (6.1) |  |
| **Obesity diagnosis history** |  |  |  | 0.002 |  |  | 0.27^†^ |
| No | 521 (85.6) | 36 (70.6) | 485 (86.9) |  | 12 (75.0) | 509 (85.8) |  |
| Yes | 88 (14.5) | 15 (29.4) | 73 (13.1) |  | 4 (25.0) | 84 (14.2) |  |
| **Arthritis diagnosis history** ^†^ |  |  |  | 0.02 |  |  | 0.05 |
| No | 553 (90.8) | 41 (80.4) | 512 (91.8) |  | 12 (75.0) | 541 (91.2) |  |
| Yes | 56 (9.2) | 10 (19.6) | 46 (8.2) |  | 4 (25.0) | 52 (8.8) |  |
| **Stroke diagnosis history** ^†^ |  |  |  | 0.17 |  |  | 0.21 |
| No | 600 (98.5) | 49 (96.1) | 551 (98.8) |  | 15 (93.8) | 585 (98.7) |  |
| Yes | 9 (1.5) | 2 (3.9) | 7 (1.3) |  | 1 (6.3) | 8 (1.4) |  |
| **Preferred position for anal intercourse** |  |  |  | 0.23 |  |  | 0.06 |
| Always/mostly insertive | 145 (23.8) | 8 (15.7) | 137 (24.6) |  | 1 (6.3) | 144 (24.3) |  |
| Versatile | 246 (40.4) | 20 (39.2) | 226 (40.5) |  | 5 (31.3) | 241 (40.6) |  |
| Always/mostly receptive | 218 (35.8) | 23 (45.1) | 195 (35.0) |  | 10 (62.5) | 208 (35.1) |  |
| **Frequency of anal intercourse in the last 30 days** |  |  |  | 0.80^⸹⸹^ |  |  | 0.05^⸹⸹^ |
| 0 | 186 (31.0) | 20 (40.0) | 166 (30.2) |  | 10 (62.5) | 176 (30.1) |  |
| 1 | 87 (14.5) | 6 (12.0) | 81 (14.7) |  | 2 (12.5) | 85 (14.6) |  |
| 2-3 | 144 (24.0) | 5 (10.0) | 139 (25.3) |  | 0 | 144 (24.7) |  |
| 1 per week | 94 (15.7) | 10 (20.0) | 84 (15.3) |  | 2 (12.5) | 92 (15.8) |  |
| > 1 per week | 89 (14.8) | 9 (18.0) | 80 (14.6) |  | 2 (12.5) | 87 (14.9) |  |
| **Last receptive anal intercourse** ^†^ |  |  |  | 0.60 |  |  | 0.40 |
| Within last month | 282 (46.6) | 23 (45.1) | 259 (46.8) |  | 5 (31.3) | 277 (47.0) |  |
| > 1 month | 276 (45.6) | 23 (45.1) | 253 (45.7) |  | 11 (68.8) | 265 (45.0) |  |
| Never | 21 (3.5) | 1 (2.0) | 20 (3.6) |  | 0 | 21 (3.6) |  |
| Don’t know | 26 (4.3) | 4 (7.8) | 22 (4.0) |  | 0 | 26 (4.4) |  |
| **Clinician type** |  |  |  | <0.001 |  |  | 0.53 |
| Medical doctor | 275 (45.2) | 9 (17.7) | 266 (47.7) |  | 6 (37.5) | 269 (45.4) |  |
| Advanced practice nurse | 334 (54.8) | 42 (82.4) | 292 (52.3) |  | 10 (62.5) | 324 (54.6) |  |
| Data are n (%). Missing: Waist size n = 4; gender identity n = 1; race and ethnicity n = 4; sexual orientation n = 1; years of school n = 5; marital n = 7; HIV diagnosis history n = 8; frequency of anal intercourse in the last 30 days n = 9; last receptive anal intercourse n = 4. Abbreviations: IQR, interquartile range; SD, standard deviation. All hypothesis tests are Pearson chi square unless otherwise noted. | | | | | | | |
| * Student’s *t*-test | | | | | | | |
| ^†^ Fisher’s Exact Test | | | | | | | |
| ^‡^ Other includes Native Hawaiian or Pacific Islander, American Indian or Alaskan Native, and Other. | | | | | | | |
| ^**^ Individuals’ self-reported sexual orientation included ‘lesbian’ (n=1) although self-reported sexual behavior satisfied inclusion criteria. | | | | | | | |
| ^⸹⸹^ Cochran-Armitage Test for Trend | | | | | | | |

| **Supplemental Table 2*:* Factors associated with sphincter resting tone measured by physicians and advanced practice nurses completing >50 digital anal rectal examinations, Chicago and Houston, USA 2020-2022** | | |
| --- | --- | --- |
|  | **Resting tone** | |
|  | **OR (95% CI)** | **aOR*** **(95% CI)** |
| **Age**, years | **1.08 (1.05-1.11)** | **1.08 (1.05-1.12)** |
| **Body mass index** | 1.03 (0.99-1.07) | - |
| **Waist size,** centimetres |  |  |
| ≤ 102 | 1.0 | - |
| > 102 | **2.34 (1.32-4.16)** | - |
| **Gender identity** |  |  |
| Man | 1.0 | - |
| Transgender woman, transgender man, woman, and other | 2.83 (0.81-9.83) | - |
| Non-binary | 0.33 (0.02-6.09) | - |
| **Sexual orientation** |  |  |
| Gay | 1.0 | - |
| Bisexual | 0.57 (0.15-2.13) | - |
| Queer | 0.17 (0.01-2.92) | - |
| Heterosexual, lesbian, other, or don’t know ** | 1.97 (0.30-12.97) | - |
| **Diabetes diagnosis history** |  |  |
| No | 1.0 | - |
| Yes | **3.27 (1.43-7.48)** | - |
| **Obesity diagnosis history** |  |  |
| No | 1.0 | 1.0 |
| Yes | **2.81 (1.47-5.35)** | **2.78 (1.37-5.63)** |
| **Arthritis diagnosis history** |  |  |
| No | 1.0 | - |
| Yes | **2.79 (1.32-5.88)** | - |
| **Preferred position for anal intercourse** |  |  |
| Always/mostly insertive | 1.0 | 1.0 |
| Versatile | 1.46 (0.64-3.36) | 1.47 (0.60-3.61) |
| Always/mostly receptive | 1.95 (0.86-4.40) | **2.52 (1.04-6.12)** |
| **Frequency of anal intercourse in the last 30 days** |  |  |
| 0 | 1.0 |  |
| 1 | 0.65 (0.26-1.64) | - |
| 2-3 | **0.32 (0.12-0.85)** | - |
| 1 per week | 1.01 (0.46-2.23) | - |
| > 1 per week | 0.96 (0.42-2.17) | - |
| **Clinician type** |  |  |
| Medical doctor | 1.0 | 1.0 |
| Advanced practice nurse | **4.08 (1.98-8.41)** | **3.07 (1.43-6.58)** |
| Before rounding, bolded point estimates and 95% CI did not include unity. Abbreviations: OR, odds ratio; CI, confidence interval; aOR, adjusted odds ratio. | | |
| * Adjusted by variables retained in model and potential confounders assigned sex at birth, race and ethnicity, and HIV. | | |

| **Supplemental Table 3*:* Accuracy of digital anal rectal examinations to detect persons engaging in any receptive anal intercourse* as measured by physicians and advanced practice nurses completing >50 examinations, Chicago, and Houston USA 2020-2022** | | |
| --- | --- | --- |
|  | **Resting tone accuracy^†^ (95% CI)** | **Squeeze tone accuracy^†^ (95% CI)** |
| **Overall** | 0.52 (0.50-0.54) | 0.51 (0.50-0.52) |
| **Clinician type** |  |  |
| Medical doctor | 0.52 (0.51-0.54) | 0.51 (0.50-0.53) |
| Advanced practice nurse | 0.52 (0.48-0.56) | 0.51 (0.50-0.53) |
| * Any receptive anal intercourse defined as a preferred position for anal intercourse that is either versatile or always/mostly receptive anal sex. | | |
| **^†^** Area under the receiver operating characteristic curve | | |
